# Real-World Usage Patterns of Large Language Models in Healthcare

**DOI:** 10.1101/2025.05.02.25326781

**Authors:** Alyssa Unell, Mehr Kashyap, Michael Pfeffer, Nigam Shah

## Abstract

**Objective:** To characterize real-world LLM use by healthcare professionals and identify gaps between actual usage and research focus.

**Materials and Methods:** We analyzed chat interactions from a secure deployment of GPT-3.5/4 at an academic medical center (Dec 2023-May 2024), classifying tasks using GPT-4o-mini according to a published taxonomy.

**Results:** Among 25,173 interactions from 3,913 users, 64.1% were healthcare-related. Most common tasks were writing for professional communication (23.9%), enhancing medical knowledge (12.5%), general writing support (9.7%), and medical research (8.7%). Note-taking (1.3%) and billing/coding (0.35%) were rare. Task frequencies correlated moderately with literature (r = 0.75).

**Discussion:** Real-world usage diverged from literature in key areas. Writing support and medical research tasks were prevalent in practice yet underexplored in research. Clinical decision support, note-taking, and billing showed limited adoption despite being promising applications, suggesting workflow and implementation barriers.

**Conclusion:** These findings help hospital administrators align LLM deployment with actual needs and address implementation barriers.

## BACKGROUND AND SIGNIFICANCE

Large language models (LLMs) can assist in a wide range of healthcare tasks, from clinical decision-making at the bedside to administrative tasks [1-6]. While research literature has explored numerous potential applications, less is known about how healthcare professionals actually utilize these tools when given access in real-world clinical settings [7-10]. This discrepancy between theoretical applications and practical usage represents a significant knowledge gap for healthcare organizations planning LLM implementations.

Our medical center provided access to a secure instance of GPT-3.5 and GPT-4 to enable safe use of LLMs for a variety of tasks potentially involving Protected Health Information (PHI)[11]. This secure environment created an opportunity to study authentic usage patterns in a healthcare setting where privacy concerns were addressed through institutional safeguards.

## OBJECTIVES

This study serves a dual purpose: (1) identifying gaps in workflow integration by analyzing actual usage patterns across different healthcare roles, and (2) comparing real-world usage with academic literature predictions to highlight potential misalignments between research focus and practical needs. By examining six months of real-world LLM usage patterns by healthcare professionals and comparing real-world usage of this technology with proposed usage by academic literature, we can identify both the most valuable current applications as well as areas where potential adoption barriers may exist for otherwise beneficial applications.

Understanding these patterns is particularly relevant for healthcare informatics departments tasked with implementing and optimizing LLM tools within complex clinical environments. This data can inform decisions about resource allocation, system design, and prioritization of integration efforts to maximize the clinical and operational value of these emerging technologies [12,13,14].

## MATERIALS AND METHODS

### System Implementation and Data Collection

We analyzed usage data from Secure GPT, a safe platform offering access to LLMs, such as GPT-3.5 and GPT-4 from OpenAI, to 33,000 healthcare and academic users at Stanford Medicine. Users included about 3,500 doctors, 5,000 nurses, 3,500 students, 2,900 postgraduate clinical and research trainees, and remaining users were other healthcare professionals.

All interactions from December 15, 2023, to May 8, 2024 (approximately six months) were extracted in de-identified form for analysis. The extraction process removed personal identifiers while preserving essential metadata including the complete conversation content necessary for classification. De-identification was performed at the user level, removing all identifiers while preserving conversation content. Examples of real user queries can be found in Table 1.

**Table 1.** We include examples of de-identified healthcare-related instructions and questions submitted by healthcare professionals on the Secure GPT platform.

|  |
| --- |
| Write a letter to my patient's employer requesting an accommodation for sufficient break time so she can take her medications. |
| How long should clopidogrel be held in patients undergoing colonoscopy? |
| Create a nursing note for a patient who is four days post-allogeneic stem cell transplant. |
| How do I inform my patient they have cancer? |
| Can you explain what Covered California is? |
| What is a good way to visualize mass spectrometry-based proteomics data? |

### Analysis Framework

We employed a multi-step approach combining automated classification with expert-level review of user-LLM interactions. Additional information regarding the prompts used for classification can be found in the supplementary materials. We used GPT-4o-mini to label interactions according to healthcare tasks from Bedi et al.’s systematic review of 519 studies evaluating LLM applications in healthcare [15]. In Bedi’s framework, each published study was categorized by its primary healthcare application, allowing us to compare the distribution of research attention across different tasks to real-world usage patterns.

Our task labeling process followed two primary steps:

1. First, GPT-4o-mini identified whether each interaction was healthcare-related.
2. For healthcare-related interactions, it classified them into one of the tasks from Bedi et al.’s taxonomy, allowing for an “other” task label when the interaction did not fit established categories.

We estimated agreement between GPT-4o-mini’s assigned tasks and a three-author consensus on a random sample of 71 healthcare-classified interactions out of an original sample of 100 interactions where 29 were discarded due to lack of relevance. For interactions labeled as “other,” we conducted a qualitative analysis to identify emerging task categories not captured in Bedi et al.’s review that may be prevalent in real world deployment. After developing these new task categories, GPT-4o-mini was then used to reclassify all interactions initially labeled as “other” into these newly identified task categories.

To assess alignment between practical usage and research literature, we compared the frequency distribution of observed tasks with their representation in published studies as reported by Bedi et al., calculating a Pearson correlation coefficient to quantify this relationship. This comparison helps identify potential misalignments between areas receiving research attention and those providing practical value, thereby informing more targeted research and implementation efforts.

## RESULTS

### Classification Consensus

The raw agreement rate between the human consensus and our model judge was 64.8%. The raw agreement rate reflects the inherent challenge of categorizing multi-purpose interactions. Given the large number of possible task categories, chance agreement between model and human judges would be approximately 4.5%. As such, our agreement rate of 64.8% indicates reasonable alignment between automated classification and human judgment. Furthermore, 84% of the questions miscategorized by our model judge were subject to additional internal discussion among the human annotators to arrive at consensus, further suggesting that the disagreements largely occurred in ambiguous cases rather than clear misclassifications. In 71% of instances where the model was incorrect and there was internal discourse, the model’s classification matched at least one human reviewer classification.

### Usage overview

We analyzed 25,173 chat interactions, encompassing 354,048 instruction-response exchanges, from 3,913 healthcare and academic users at a single medical center over the six-month period. Of the total interactions, 16,137 (64.1%) were classified as healthcare-related.

Among healthcare-related interactions, 7,539 (46.7%) corresponded to the healthcare tasks described in Bedi et al., while the remainder (53.3%) were initially classified as “other” and required new task classifications to be developed through manual review. Qualitative analysis of interactions initially labeled as “other” revealed several prevalent tasks not explicitly captured in the Bedi et al. framework, including Supporting Career Development, Generating Performance Evaluations, Writing Assistance for Professional Communication, and General Writing Support (Table 2).

**Table 2.** Real-world usage patterns of large language models by healthcare professionals, comparing chat interactions by task with existing literature. ⋆ The total number and percentage of interactions labeled as ‘Other’ are shown in parentheses, with specific tasks provided within this category.

| **Healthcare Task** | **No. of chat interactions** | **Percentage of chat interactions** | **Percentage of chat interactions (Bedi et al. tasks only)** | **Percentage of studies in literature (Bedi et al.)** |
| --- | --- | --- | --- | --- |
| *Biomedical data mining* | 11 | 0.07% | 0.15% | 0.30% |
| *Care coordination and planning* | 387 | 2.40% | 5.13% | 5.90% |
| *Carrying out a literature review* | 1028 | 6.37% | 13.64% | 2.90% |
| *Clinical note-taking* | 216 | 1.34% | 2.87% | 0.60% |
| *Communicating with patients* | 353 | 2.19% | 4.68% | 5.90% |
| *Conducting medical research* | 1,398 | 8.66% | 18.54% | 1.30% |
| *Educating patients* | 679 | 4.21% | 9.01% | 14.00% |
| *Enhancing medical knowledge* | 2,009 | 12.45% | 26.65% | 35.20% |
| *Enhancing surgical operations* | 9 | 0.06% | 0.12% | 0.50% |
| *Generating clinical referrals* | 17 | 0.11% | 0.23% | 0.20% |
| *Generating medical reports* | 152 | 0.94% | 2.02% | 1.30% |
| *Generating provider billing codes* | 56 | 0.35% | 0.74% | 0.50% |
| *Making diagnoses* | 688 | 4.26% | 9.13% | 15.40% |
| *Making treatment recommendations* | 485 | 3.01% | 6.43% | 7.30% |
| *Managing clinical knowledge* | 9 | 0.06% | 0.12% | 1.00% |
| *Providing asynchronous care* | 0 | 0.00% | 0.00% | 1.20% |
| *Synthesizing data for research* | 21 | 0.13% | 0.28% | 2.60% |
| *Triaging patients* | 6 | 0.04% | 0.08% | 3.60% |
| *Writing prescriptions* | 15 | 0.09% | 0.20% | 0.20% |
| *Other\** | (8598) | (53.28%) | (--) | (--) |
| *Supporting career development* | 324 | 2.01% | -- | -- |
| *Generating performance evaluations* | 201 | 1.25% | -- | -- |
| *Writing assistance for professional communication* | 3856 | 23.90% | -- | -- |
| *General writing support* | 1557 | 9.65% | -- | -- |
| *Not categorized* | 2660 | 16.48% | -- | -- |
| **Total** | 16137 | 100.00% | 100.00% | 100.00% |

### Task Distribution

The most frequent tasks were writing assistance for professional communication (23.90%, including drafting emails, reports, and other professional communications), enhancing medical knowledge (12.45%, involving questions seeking clinical information or explanation of medical concepts), general writing support (9.65%, involving assistance in writing grants and public notifications), and conducting medical research (8.66%, such as writing research papers or conducting original research).

### Comparison with Published Literature

While task frequency in real-world usage showed moderate correlation with task frequency in the literature (Pearson r=0.75), several prevalent tasks were under-reported or absent in published studies. Writing assistance for professional communication constituted nearly a third of real-world usage but represented a much smaller proportion of published studies. Similarly, tasks related to career development, performance evaluations, conducting medical research, and literature reviews showed significant disparities between practical usage and research focus. Figure 1 illustrates this misalignment between practical application and research priorities.

**Figure 1.**
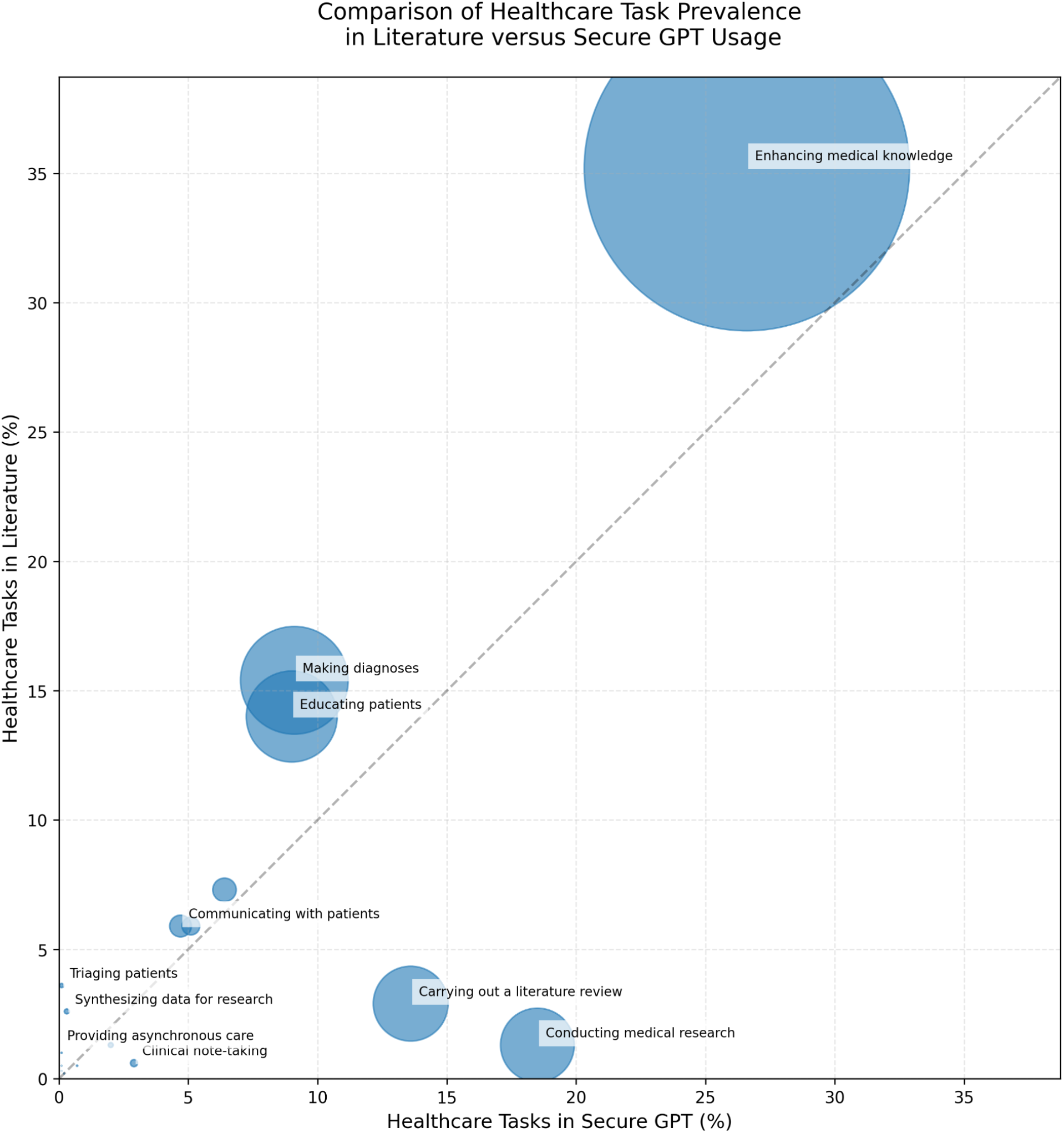
Comparison of prevalence of healthcare tasks studied in the literature versus observed in Secure GPT usage. * Circle size indicates product of the healthcare task prevalence in literature and Secure GPT usage

### Challenges with Workflow Integration

A number of proposed applications for LLMs in healthcare showed relatively low adoption in practice. Clinical decision support applications such as making diagnoses and making treatment recommendations, which constitute a substantial portion of the literature, represented a smaller fraction of combined real-world usage. Similarly, automated documentation and medical coding showed lower practical adoption than their research prominence would suggest, especially in light of new lines of work that focus on clinical note generation [16, 17]. Tasks such as clinical note-taking (1.34%) and billing and coding (0.35%) appeared infrequently in our log analysis, although these administrative areas are key applications that are proposed to increase hospital efficiency with LLMs. These findings suggest that technical capabilities alone are not the only consideration for adoption; successful integration requires careful attention to workflow integration, verification processes, and alignment with existing clinical systems.

## DISCUSSION

### Alignment Between Research and Practice

Healthcare and academic users at a medical center showed sustained usage of LLMs for medically relevant tasks. Our findings identify both the correlation and gaps between the focus of academic research on LLMs in healthcare and their actual usage patterns in a clinical setting. The moderate correlation suggests that while research generally tracks practical needs, there are discrepancies in specific areas. One example includes the increased prevalence of writing/communication assistance in real world usage compared to research literature. This suggests that healthcare professionals find significant value in LLMs for crafting emails, reports, and other professional communications—a use case that has received relatively little formal research attention. As such, integration of these models with communication tools and templates could provide significant value to healthcare professionals.

Enhancing medical knowledge was a commonly seen task in both research and practice, with more research focus being put on model ability to answer medical exam-like questions than was seen in practice. This indicates that researchers need to be exploring clinical model performance beyond multiple choice exams. There are many additional tasks that are underrepresented in practice and are an indication that there exists systematic barriers to the usage of these tools in an efficient way, such as automated documentation and medical coding.

### Study Limitations

This study has several limitations that should be considered when interpreting the results. First, as a single-institution study at an academic medical center with unique technological infrastructure and organizational culture, our findings may not generalize to other healthcare settings with different organizational structures, specialties, and technological resources. Second, our task labeling approach may not fully capture multifaceted interactions where users employ LLMs for multiple purposes within a single conversation. Our use of GPT-4o-mini for classification, while efficient for analyzing large datasets, introduces potential biases in how interactions are categorized. While our classification achieved moderate alignment with human judgment, the ambiguity in categorizing certain interactions highlights the challenge of developing precise taxonomies for the diverse ways healthcare professionals use these tools. Third, this study focused on usage patterns without assessing the quality, accuracy, or clinical impact of LLM responses. Understanding these dimensions is crucial for evaluating the overall utility and safety of LLMs in healthcare settings and requires separate dedicated investigation. Finally, our analysis captures an early adoption phase of LLM technology in healthcare. Usage patterns may evolve as users become more sophisticated in their interactions with these systems and as the technology itself continues to advance.

## CONCLUSION

This study presents a quantitative analysis of how healthcare professionals used LLMs in practice when offered in a secure setting over six months. The dual-purpose analysis reveals both the actual usage patterns in a healthcare environment and the alignment between these patterns and academic literature. Our findings highlight several key insights for healthcare informatics:

- A significant disconnect exists between some highly researched applications and their practical adoption, suggesting potential technical or workflow considerations that may benefit from further investigation as well as encouraging more realistic benchmarking efforts from the clinical AI community.
- Healthcare professionals find substantial value in LLM applications that have received relatively little research attention, particularly for communication tasks, research support, and professional development.
- Successful implementation likely requires both technical capabilities and careful attention to workflow integration, verification processes, and alignment with existing systems.

Future research should explore specific factors affecting adoption for heavily studied but underutilized applications, evaluate the quality and impact of LLM responses across different task categories, and investigate how different healthcare settings and specialties may show varying usage patterns and needs.

As healthcare organizations continue to explore and implement LLMs, understanding these real-world usage patterns can help guide resource allocation, system design, and integration efforts to maximize the value of these emerging technologies for healthcare professionals and the patients they serve.

## Data Availability

All data was deidentified and available within secure Stanford ecosystem.

## Notes

### Competing Interest Statement

Dr. Shah reported being a cofounder of Prealize Health (a predictive analytics company) and Atropos Health (an on-demand evidence generation company); receiving funding from the Gordon and Betty Moore Foundation for developing virtual model deployments; and serving on the Board of the Coalition for Healthcare AI (CHAI), a consensus-building organization providing guidelines for the responsible use of artificial intelligence in health care. Dr. Shah serves as a scientific advisor to Opala, Curai Health, Arsenal Capital and JnJ Innovative Medicines.

### Funding Statement

The study was funded by the Stanford Graduate Fellowship and NSF GRFP.

### Author Declarations

This is anonymized data and hence exempt (not human subjects research).

### Summary of Updates

Acknowledgement of joint first-authorship between Unell and Kashyap

