## Supplemental Data for "Real-World Usage Patterns of Large Language Models in Healthcare"

### Secure GPT Chat Interaction Labeling Process

We used GPT-4o-mini to label all Secure GPT interactions, using the following classification process. First, each interaction was labeled as either healthcare-related or non-healthcare-related. Among those labeled as healthcare-related, we further classified each interaction according to specific healthcare tasks outlined in Bedi et al, allowing for an “other” task label. Finally, all interactions labeled as "other" were reclassified into new healthcare tasks categories. The following prompts were used for this labeling process:

<u>Prompt for identifying healthcare-related Secure GPT interactions:</u>

> *Is the following question/conversation related to healthcare or medicine? Return 0 if it is healthcare related, return 1 if it is not healthcare related.*

<u>Prompt for classifying Secure GPT interactions by healthcare task:</u>

> *Given the following questions from a healthcare professional to a language model, please classify the questions as one of the following categories. Output a string of the category that you assign to the question, with the category either being a category from Category Options or a new string category. Do not provide an explanation or any other text besides the category name. Here are the category options as well as a definition and an example for each option:*
>
> *Healthcare Task: Enhancing medical knowledge*
> *Definition: The process of enhancing the skills, knowledge, and capabilities of healthcare professionals to meet the evolving needs of healthcare delivery.*

*Example: Measuring the performance of GPT on Neurosurgery Written Board examinations*
*Additional instruction: This includes requesting more information about a healthcare topic for the purpose of learning*

*Healthcare Task: Making diagnoses*
*Definition: The process of identifying the nature or cause of a disease or condition through the examination of symptoms, medical history, and diagnostic tests.*
*Example: Comparing the performance of GPT and Physicians for diagnostic accuracy*

*Healthcare Task: Educating patients*
*Definition: Providing patients with information and resources to help them understand their health conditions, treatment options, etc. for more informed decision-making around their care.*
*Example: Using GPT for Patient Information in periodontology*

*Healthcare Task: Making treatment recommendations*
*Definition: The process of providing treatment recommendations for patients to manage or cure their health conditions.*
*Example: Using GPT for therapy recommendations in mental health*

*Healthcare Task: Communicating with patients*
*Definition: The exchange of information from healthcare providers to patients. This could be done via patient messaging platforms, or via Chatbots integrated into the provider workflow.*
*Example: Using GPT to communicate with palliative care patients*

*Healthcare Task: Care coordination and planning*

*Definition: The process of organizing and integrating healthcare services to ensure that patients receive the right care at the right time, involving communication and collaboration.*

*Example: Measuring the reliability and quality of nursing care planning generated*

*Healthcare Task: Triaging patients*

*Definition: Clinical triage is the process of prioritizing patients based on the severity of their condition and the urgency of their need for care.*

*Example: Measuring the accuracy of patient triage in parasitology examination*

*Healthcare Task: Carrying out a literature review*

*Definition: A literature review is a critical summary and evaluation of existing research or literature on a specific topic.*

*Example: Examining the validity of ChatGPT in identifying relevant Nephrology literature*

*Healthcare Task: Synthesizing data for research*

*Definition: Data synthesis refers to the process of combining and analyzing data from multiple sources to generate new insights, draw conclusions, or develop a comprehensive understanding of a topic.*

*Example: Synthesizing radiologic data for effective clinical decision-making*

*Healthcare Task: Generating clinical referrals*

*Definition: A referral is an order that a medical provider places to send their patient to a specialized physician or department for further evaluation, diagnosis, or treatment.*

*Example: Assistance in optimizing Emergency Department radiology referrals and imaging selection*

*Healthcare Task: Generating medical reports*
*Definition: An image-captioning task of producing a professional report according to input image data.*
*Example: Assessing the feasibility and acceptability of ChatGPT generated radiology report summaries for cancer patients*

*Healthcare Task: Managing clinical knowledge*
*Definition: The process of ensuring clinical knowledge bases is correct, consistent, complete, and current.*
*Example: Using GPT models for phenotype concept recognition*

*Healthcare Task: Providing asynchronous care*
*Definition: A proactive way to ensure that everyone assigned to a clinic is up to date on basic preventive care, like cancer screenings or immunizations, and that they receive extra help if they have lab numbers that are high.*
*Example: Asynchronously answering patient questions pertaining to erectile dysfunction*

*Healthcare Task: Clinical note-taking*
*Definition: The process of recording detailed information about a patient's health status, medical history, symptoms, physical examination findings, diagnostic test results, and treatment plans, typically documented in the patient's EMR.*
*Example: Using GPT models for taking notes during primary care visits*

*Healthcare Task: Enhancing surgical operations*

*Definition: The process of supporting healthcare professionals, such as surgical technologists, nurses, and other staff, during surgical procedures.*

*Example: Using GPT to pinpoint innovations for future advancements in general surgery*

*Healthcare Task: Conducting medical research*

*Definition: Definition: Medical research generation, including writing papers, refers to the process of conducting original research in medicine or healthcare and documenting the findings in academic papers.*

*Example: Using GPT models for sentiment analysis of COVID-19 survey data*

*Additional instruction: Conducting medical research includes requests for assistance with coding for the purpose of data analysis (e.g. using tools such as R, SQL, etc.)*

*Healthcare Task: Biomedical data mining*

*Definition: The process of searching and extracting data regarding a patient's health.*

*Example: Using GPT models to mine and generate biomedical text (Chen et al)37*

*Healthcare Task: Generating provider billing codes*

*Definition: Medical billing is the process of submitting and following up on claims with health insurance companies to receive payment for healthcare services provided to patients.*

*Example: Using GPT models to predict diagnosis-related group (DRG) codes for hospitalized patients*

*Healthcare Task: Writing prescriptions*

*Definition: The process by which a healthcare provider, typically a physician or other qualified medical professional, orders medications or treatments for a patient.*
*Example: Prescription of kidney stone prevention treatment*

*Healthcare Task: Interprofessional communication*
*Definition: Exchanges between healthcare professionals and staff across various roles within the system, excluding direct patient involvement*
*Example: Help me improve writing for or rephrase this email*

*Healthcare Task: Other*
*Definition: Any tasks that fit in either none of or multiple categories*

Of note, we included 'interprofessional communication' as an additional category in the healthcare task classification prompt given its frequent occurrence in a preliminary manual analysis of a subset of interactions on Secure GPT, although this was not among the tasks described in Bedi et al.

<u>Prompt for Classifying "Other"-labeled Secure GPT interactions:</u>

*Given the following questions from a healthcare professional to a language model, please classify the questions as one of the following categories. Output a string of the category that you assign to the question, with the category either being a category from Category Options or a new string category. Do not provide an explanation or any other text besides the category name. Here are the category options as well as a definition and an example for each option.*

*Healthcare Task: Writing assistance for professional communication*

*Definition: Supporting communication between healthcare professionals across various teams*

*Example: Help me write or rephrase this email*

*Healthcare Task: Generating performance evaluations*

*Definition: The process of reviewing performance of healthcare professionals and trainees*

*Example: Assessing and conveying strengths and weaknesses of an employee*

*Healthcare Task: Supporting career development*

*Definition: The process of helping healthcare professionals with career advancement*

*Example: Creating a letter of recommendation for a junior faculty member*

*Healthcare Task: General writing support*

*Definition: Assistance with writing tasks not specifically related to professional communication*

*Example: Help me write a newsletter for our department*

*Healthcare Task: Other*

*Definition: Any tasks that fit in either none of or multiple categories*
